# A Study of a Stochastic Model and Extinction Phenomenon of Meningitis Epidemic

**DOI:** 10.1101/2024.05.04.24306871

**Authors:** S.J. Yaga, F.W.O. Saporu

## Abstract

A stochastic version of the deterministic model for meningitis epidemic by Yaga and Saporu (2024) is developed. The stochastic mean system of equations for possible state of an individual in the model and the extinction probabilities for carrier and infective are derived. Comparison of the system of stochastic mean equations and its deterministic analogue of profiles for the various compartments and the case-carrier trajectories show similar pattern with a time shift difference. This indicates that there must be caution in using the deterministic analogue as an approximating system of the stochastic mean equations for inferential purpose. Simulation studies of the comparison of the compartmental profiles for the general case; model I, with the assumption that a proportion (*φ ≠* 0), of the infected susceptible can move directly to the infective stage and that of the special case, model II, when *φ* = 0 is examined for various values of *ϵ*(odds in favour of a carrier transmitting infection) *≤* 2. It is only when *ϵ* = 2 that model II can approximate model I in all compartments except that of the carrier. Transmission rate, *β*, loss of carriership rate, *σ* and *ϵ* are identified as the most sensitive parameters of the extinction probabilities. Threshold results derived for carrier and infective extinction probabilities are distinct but bear some relation, transmission rate required for carrier extinction is square of that for infective. It is concluded that carriership play a more prominent role in the transmission of meningitis epidemic and efforts aimed at control should be targeted at reducing the transmission rate and increasing the loss of carriership.

## 1. Introduction

Meningitis is an infection of the meninges, a thin lining surrounding the brain and the spinal cord (WHO, 2021). The disease can be caused by many pathogens (bacteria, fungi or viruses). Susceptible individuals acquire pathogen after exposure through effective and prolonged contact with asymptomatic carriers or infectious individuals (Meyer and Kristiansen, 2016). Meningococcal disease occurs worldwide as an endemic devastating disease with seasonal fluctuations (Stephen et al. 2007 and Caugant et al. 2012). The risk of meningitis varies with age. So also the carrier prevalence is age-dependent (Campagne et al. 1999). This disease poses a major public health threat.

Deterministic models have been developed to study the dynamics of meningitis for endemic situations (Irving et al., 2012; Coen et al., 2000; Veren, 2008; Karachaliu et al., 2015; Asamao et al., 2018; Agier et al., 2013). Yaga and Saporu (2024) extended the work of Irving et al. (2012) for an epidemic situation using a compartmental deterministic model.

One of the assumptions of deterministic models is that the sizes of the compartments are large enough to enable homogeneous mixing in the population (Bailey, 1975 and Darley and Gani, 1999). Such a notion is not generally applicable when studying the dynamic behavior of biological populations such as diseases, population growth, and human behavior, etc. Indeed human population is affected by demographic fluctuations, environment, climate, etc. Such variable factors induce stochasticity in the population structure and pathogen characteristics. Consequently, stochastic models are useful in studying any disease transmission dynamics.

In meningitis, contact of susceptible with either infectives or carriers is assumed to be random. This provides the scope for incorporating a stochastic effect in the model for a better understanding of the transmission process. The first stochastic model for meningitis is by Stollenwerk et al. (2004). The model investigates the effect of multi-strain meningococcal pathogens on the outbreak of meningococcal meningitis. Here a stochastic model is developed from the deterministic model of Yaga and Saporu (2024) with interest in studying the salient features of the meningitis epidemic process. In particular, we are looking at;

a. comparing the profiles of the derived system of equations for the stochastic mean and its deterministic analogue,
b. comparing the simulated stochastic model and its deterministic analogue for some important epidemiological parameters that reflect the salient features of the transmission process and
c. deriving the extinction probability arising from the stochastic model and examining the parameters of the model that are sensitive to the extinction phenomenon.

All these are new.

## 2. Stochastic Model

The salient features in the transmission of meningitis epidemic has been captured using a deterministic model in Yaga and Saporu (2024). Here interest lies in its stochastic version. Consequently, for the ease of the reader, the deterministic model formulation is presented below.

Only six(6) states are used to classify an individual so that the model is simple and mathematically tractable. Consequently, an individual can be either susceptible (*S*), asymptomatic carrier (*C*), symptomatic infectious or ill (*I*), infectious with complication (*I*_1_), recovered/immune (*R*) or dead (*D*) due only to illness of the disease. Death can only occur if an individual passes through the complication stage without recovery. A susceptible individual can be infected either by a carrier (*C*) or an infectious individual (*I*) with transmission rate *β*. In order to reflect a differential transmission rate, a force of infection,

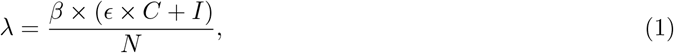

is assumed, where *N* is the total population size and *ϵ* is the odds in favor of a carrier transmitting infection to a susceptible. It is assumed that all infected individuals must pass through carriership before becoming an infective in stage *I* and infectives in stage *I*_1_ are out of circulation and, as such, do not contribute to the spread of the disease.

Carriers can either develop invasive disease (*I*) at the rate, *φ* or lose carriage at the rate, *σ* to become susceptible again. An infective in state *I* either progresses to state *I*_1_ at the rate, *θ* or recovers at the rate, *γ*_1_. An infective in state *I*_1_ either dies at the rate, *δ* or recovers at the rate, *γ*_2_. Recovered individuals lose temporary immunity to become susceptible at the rate, *α*.

In order to generalize the model, a proportion *φ* of infectives is allowed to pass directly from *S* to *I*. This reflects some of the thinking of the model of transmission of infection between infected and susceptible individuals. Finally, the model is assumed to be closed to birth, mortality and migration (in or out). For clarity, the model is diagrammatically shown in Figure <u>10</u> below.

A stochastic model is of interest. Let *S*(*t*), *C*(*t*), *I*(*t*), *I*_1_(*t*), *R*(*t*) and *D*(*t*) denote the number of susceptible, carrier, stage one infective (with symptoms of invasive disease only), infective with stage one complication, recovered and dead present at time *t*. The model implies that there are nine (9) possible transitions in Δ*t* requiring corresponding transition probabilities. For example, if one susceptible becomes a carrier in time Δ*t*, then a transition of (*S, C, I, I*_1_, *R, D*) *→* (*S −* 1, *C* + 1, *I, I*_1_, *R, D*) has occurred with transition probability given by (1 *− φ*)*β ×* (*I* + *ϵ × C*) *× N*^*−*1^(Δ*t*). Here the transition rate is denoted by *f*_*−*1,1_ = (1 *− φ*)*β ×* (*I* + *ϵ × C*) *× N*^*−*1^. By similar reasoning, the table for the transition probabilities for the stochastic model is provided in Table 1 below, where *f*_*j,k*_, *j* and *k* can take possible values *−*1, 0, 1. The possible range of parameter values are provided in Table 2.

**Tab. 1:** Showing transition probabilities between epidemiological classes.

| Description of possible events | Transition in time $t \rightarrow t + \Delta t$ | Transition Probability ($f_{jk}\Delta t$) |
| --- | --- | --- |
| 1 $S \rightarrow C$ | $(S, C, I, I_1, R, D) \rightarrow (S-1, C+1, I, I_1, R, D)$ | $(1-\varphi) \times \beta \times S \times (I + \epsilon \times C) \times \left(\frac{1}{N}\right) \Delta\tau + o(\Delta\tau)$ |
| 2 $S \rightarrow I$ | $(S, C, I, I_1, R, D) \rightarrow (S-1, C, I+1, I_1, R, D)$ | $\varphi \times \beta \times S \times (I + \epsilon \times C) \times \left(\frac{1}{N}\right) \Delta\tau + o(\Delta\tau)$ |
| 3 $C \rightarrow S$ | $(S, C, I, I_1, R, D) \rightarrow (S+1, C-1, I, I_1, R, D)$ | $\sigma \times C \Delta\tau + o(\Delta\tau)$ |
| 4 $C \rightarrow I$ | $(S, C, I, I_1, R, D) \rightarrow (S, C-1, I+1, I_1, R, D)$ | $\phi \times C \Delta\tau + o(\Delta\tau)$ |
| 5 $I \rightarrow I_1$ | $(S, C, I, I_1, R, D) \rightarrow (S, C, I-1, I_1+1, R, D)$ | $\theta \times I \Delta\tau + o(\Delta\tau)$ |
| 6 $I \rightarrow R$ | $(S, C, I, I_1, R, D) \rightarrow (S, C, I-1, I_1, R+1, D)$ | $\gamma_1 \times I \Delta\tau + o(\Delta\tau)$ |
| 7 $I_1 \rightarrow R$ | $(S, C, I, I_1, R, D) \rightarrow (S, C, I, I_1-1, R+1, D)$ | $\gamma_2 \times I_1 \Delta\tau + o(\Delta\tau)$ |
| 8 $I_1 \rightarrow D$ | $(S, C, I, I_1, R, D) \rightarrow (S, C, I, I_1-1, R, D+1)$ | $\delta \times I_1 \Delta\tau + o(\Delta\tau)$ |
| 9 $R \rightarrow S$ | $(S, C, I, I_1, R, D) \rightarrow (S+1, C, I, I_1, R-1, D)$ | $\alpha \times R \Delta\tau + o(\Delta\tau)$ |
| 10 No Change | $(S, C, I, I_1, R, D) \rightarrow (S, C, I, I_1, R, D)$ | (1-sum of all transition probabilities 1 to 9) |

**Tab. 2:**
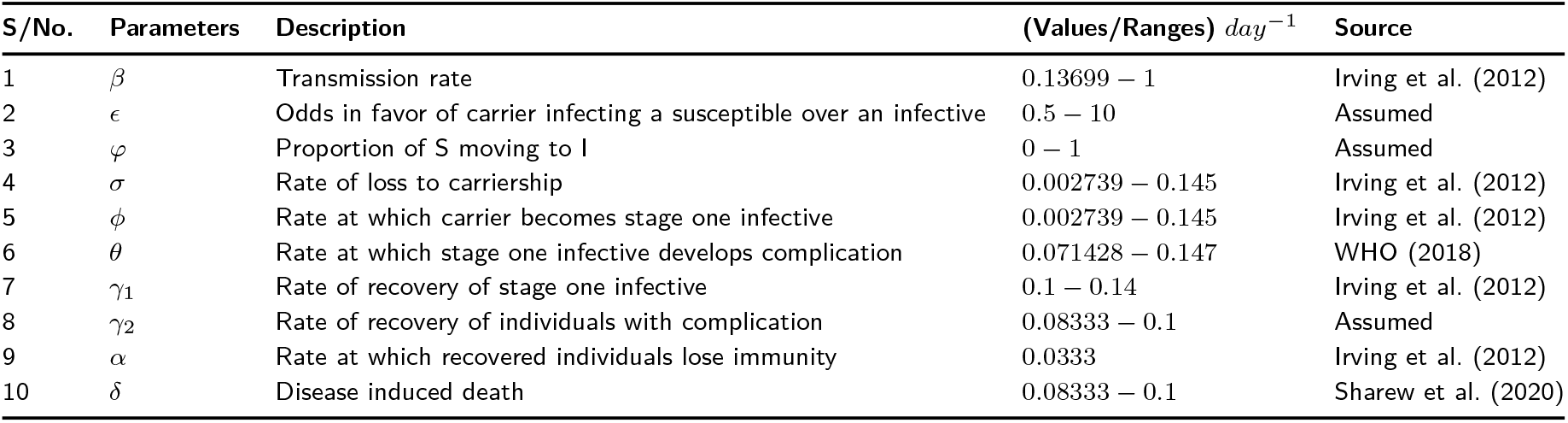
Description of parameter values used in the model and their possible range of values.

Let 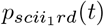 be the probability that at time *t* there are still *s* susceptibles remaining uninfected, *c* carriers in circulation, *i* infective in circulation, *i*_1_ infective with complication, *r* recovered and *d* death. By following the usual arguments by Bailey 1964, the Kolmogorov system of forward differential equations can be derived from the transition probabilities in Table 1 and are given by

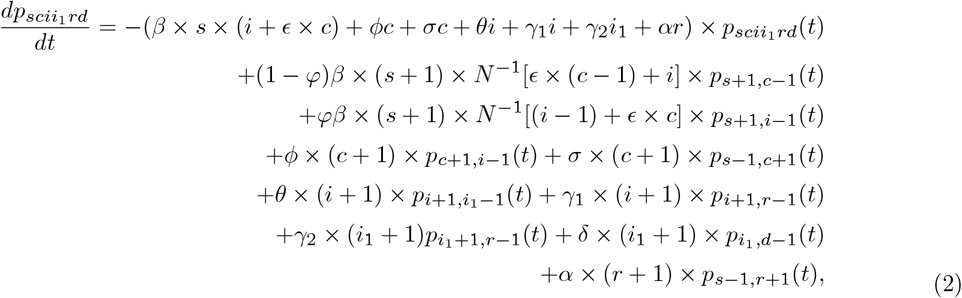

with boundary conditions *p*_*ijklmn*_(0) = 1 and initial values, *S*(0) = *n − a*_1_ *− a*_2_, *C*(0) = *a*_1_, *I*(0) = *a*_2_, *I*_1_(0) = 0, *R*(0) = 0 and *D*(0) = 0.

### 2.1. Moment Generating Function

From Table 1, the partial differential equation for the joint moment generating function *M* (*μ*_1_, *μ*_2_, *μ*_3_, *μ*_4_, *μ*_5_, *μ*_6_; *t*) can be immediately written down (following, for example, the method of Bailey, 1964, section 7.4) as

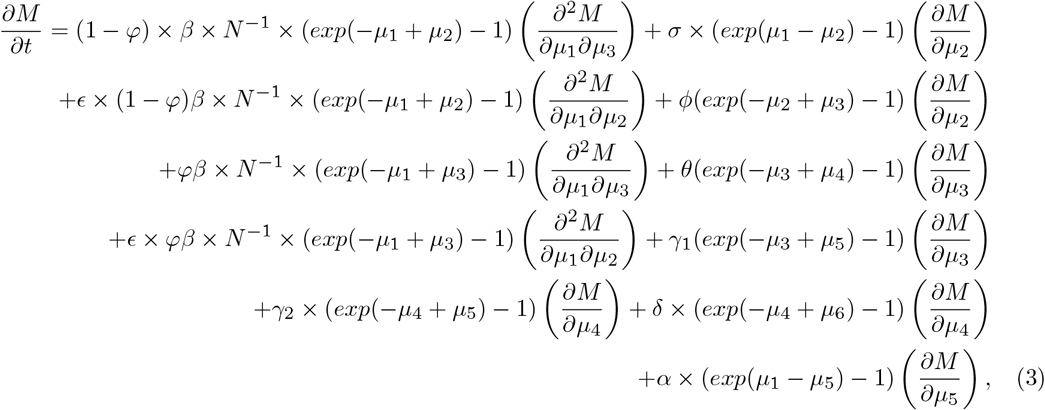

with initial condition,

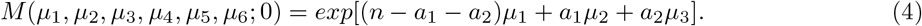

Equation (3) is intractable. However, we can obtain the system of equation for the joint moments by equating coefficients on both sides of equation (3). Let *M*_*ijklmn*_ be the (*ijklmn*)^*th*^ ordered joint moment at time *t*. Taking the procedure of equating coefficients of both sides of equation (3) as far as the second order moment, we can obtain the following equations,

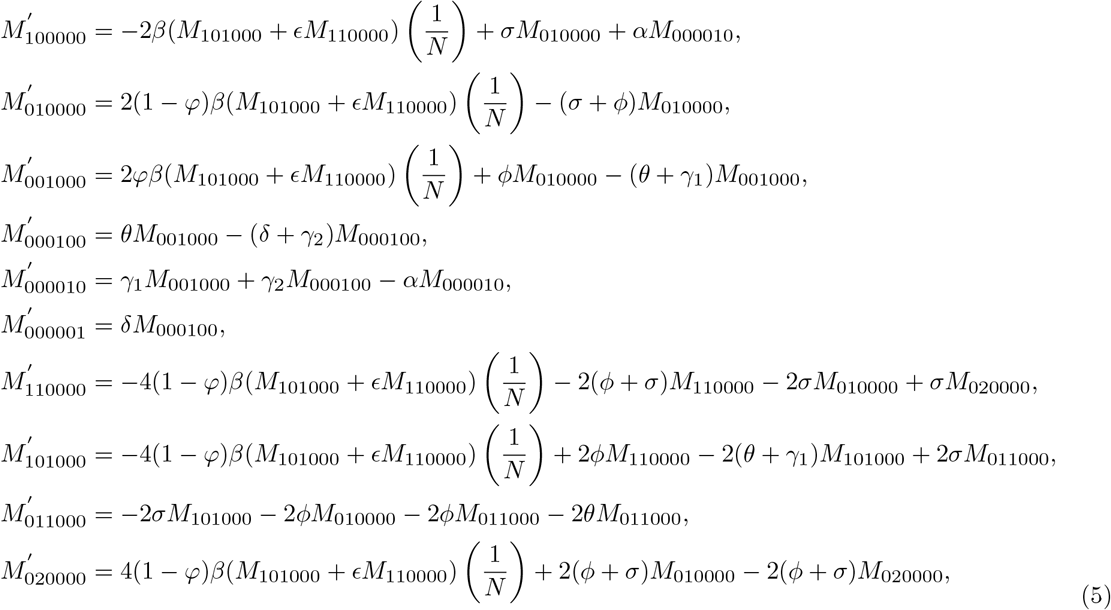

with initial conditions when *t* = 0,

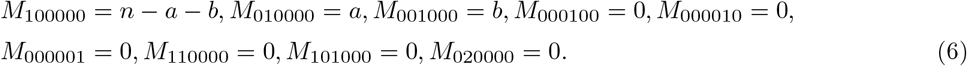

The system of equation in (5) provides the stochastic mean. It is therefore interesting to see how they compare with the deterministic system of equation given below.

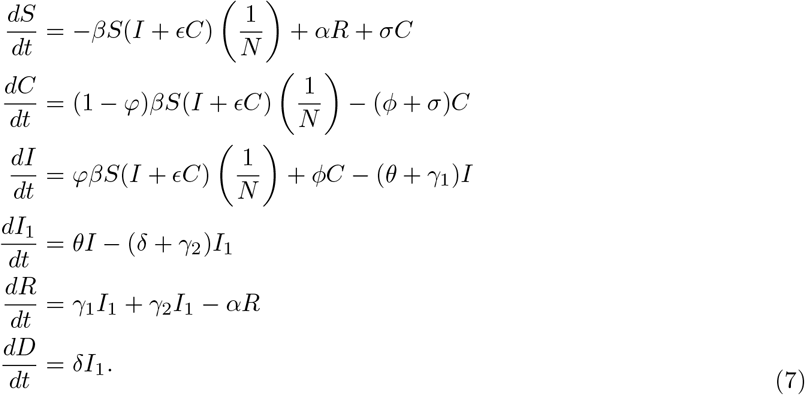

### 2.2. Comparison of the stochastic mean equations and the deterministic model profiles for meningitis epidemic process

A deterministic model in a general sense is widely used in modeling the transmission dynamics of infectious diseases and assessing the impact of various control strategies. The deterministic solution does not give an exact behavior of the corresponding stochastic mean except if the covariance between the variables is zero (Isham, 1993; Lloyd and Zhang, 2007; and Keeling and Rohani, 2008). The assumption of the meningitis model does not allow the covariance between the variables (epidemiological classes) to be zero.

Interest lies in comparing the stochastic mean and the deterministic curve in three instances to see to what extent inference from the deterministic model is dented. This will provide the necessary caution in assuming that the deterministic model adequately approximates the stochastic model for meningitis epidemic process. The first comparison is between the stochastic means and the deterministic curves of various epidemiological classes assumed for the meningitis epidemic process. The comparison is performed for population sizes *N* = 10_2_, 10^4^, 10_5_ and 10_6_ and shown in Figure 2 below. The stochastic mean equation contains joint moments that cannot be assumed to be zero, these relationship makes the stochastic mean equations slightly differ from the deterministic solution. This can explain the differences associated with the stochastic and deterministic plots. However, the nature of the pattern exhibited by both the stochastic means and the deterministic curves for all population sizes examined are the same. These provide evidence for caution in using the deterministic model as a first approximation to the stochastic mean equations.

**Fig. 1:**
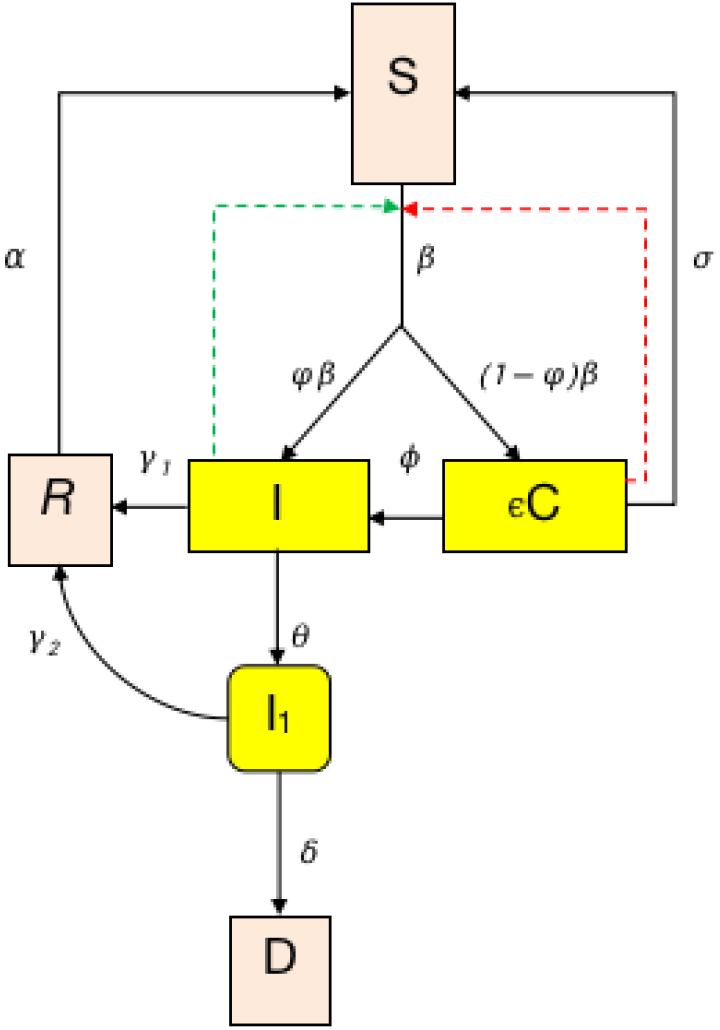
Shematic representation of the meningitis epidemic model.

**Fig. 2:**
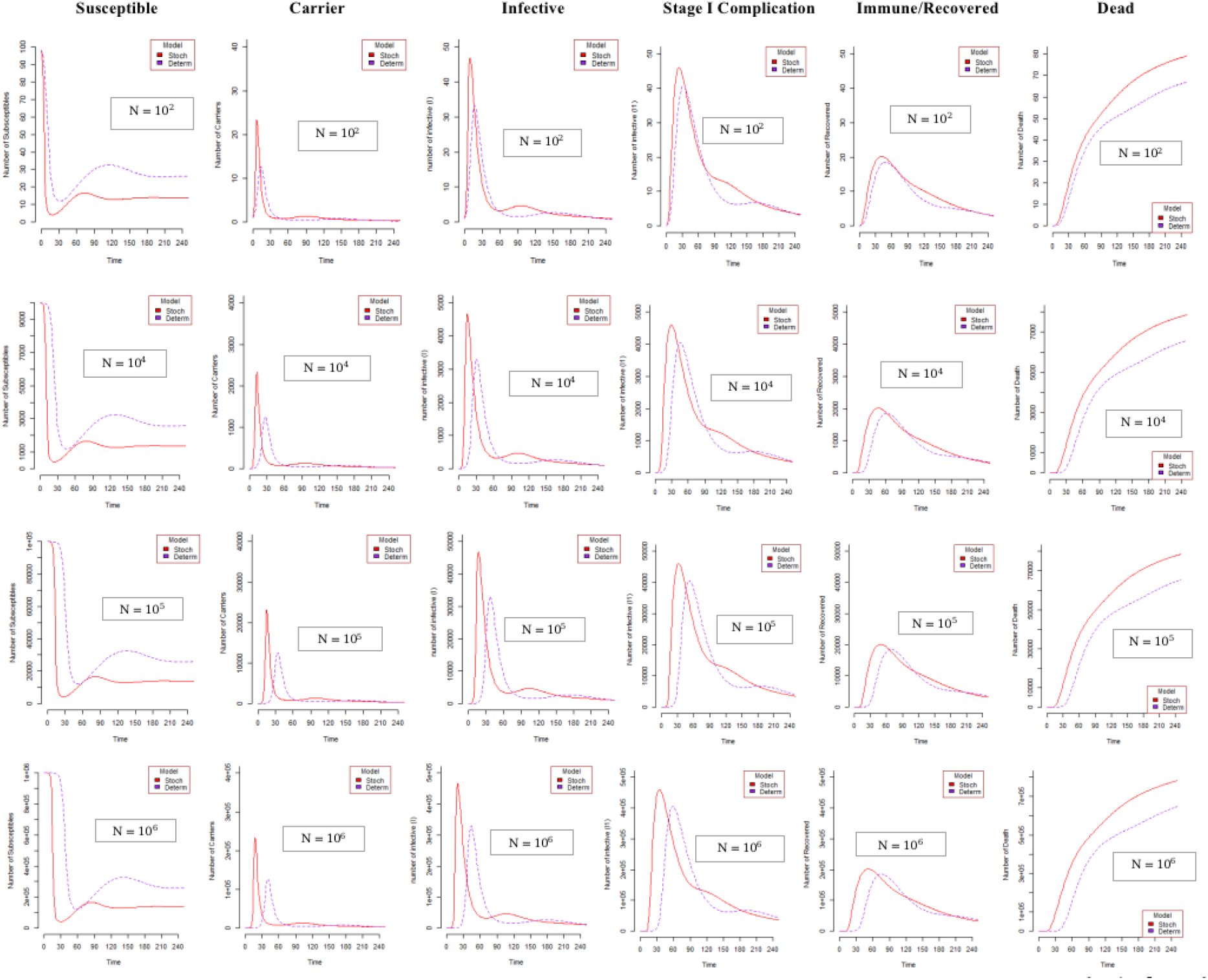
Comparison between stochastic means and deterministic model profiles for various population sizes *N* = 10^2^, 10^3^, 10^4^ and 10^6^ and parameters *ϵ* = 2 and *φ* = 0.5.

The second set of plots shown in Figure 3 below are plots of the stochastic means and deterministic profiles of carriers and infective over time for model I(*φ ≠* 0) and model II(*φ* = 0) for various initial conditions (*I*(0) = 0 and *C*(0) = 1, *I*(0) = 1 and *C*(0) = 0 and *I*(0) = 1 and *C*(0) = 1). Here, we are investigating how *φ* (the proportion of infectives that pass directly from *S* to *I*) affects these means. It is clear from all the graphs that there is covariability between the numbers of carrier and infectives over time. This gives credence to the observable differences in the stochastic means and the deterministic plots shown in Figure 2. It is noticeable from Figure 3 that;

**Fig. 3:**
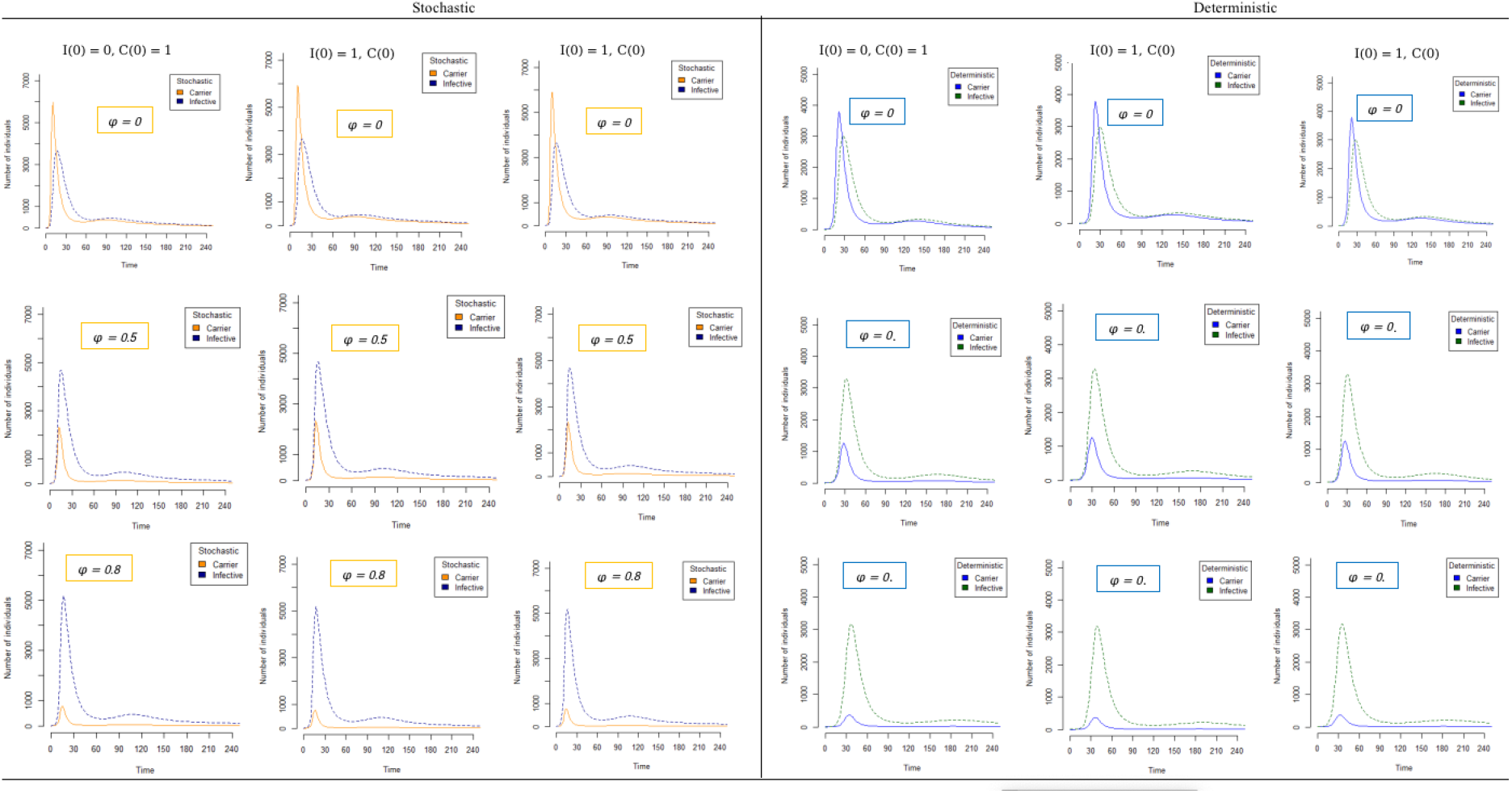
Plots of the stochastic means and the deterministic profiles for carrier and infective for model I(*φ ≠* 0) and model II (*φ* = 0)

1. the peak values of the profiles for the deterministic model underestimate the corresponding values for the stochastic mean and
2. the distinction between model I and II becomes clearer as *φ* moves further away from zero for both the deterministic and stochastic mean profiles.

Consequently, model I can approximate model II only for values of *φ* very close to zero. In such a case, the deterministic model can be used, to approximate the stochastic model bearing in mind we are contending with underestimation.

Lastly, we compared the case-carrier ratio trajectories for the stochastic means and the deterministic model. It should be mentioned here that the case-carrier ratio is an ecological proxy for the risk of meningitis giving colonization. The importance of case-carrier ratio trajectory is in providing visual evidence of how meningitis incidence varies according to epidemiological context (endemicity, hyper-endemicity, and epidemic) times. These are extensively discussed in Yaga and Saporu (2024). This informed the choice for this comparison. We are again looking to see if the deterministic model equations can serve as a first approximation to the stochastic mean equations for inference purposes.

Figure 4 shows the trajectories of case-carrier ratio for the stochastic means and deterministic model for various initial values as indicated. It must be noted that the threshold line is provided in the graph because it divides the graph into two epidemic regions (above the line is the non-epidemic region and below the line is the epidemic region) for ease of interpretation by the reader. The plots for both the stochastic mean and deterministic model show the same pattern for all the cases considered. However, plots for the deterministic model are shifted in time a little bit to the right. This implies that there is a delay in the deterministic cases in the times recorded for important epidemiological events. For examples, the times the epidemic starts, finishes, and settles in an endemic level before the next epidemic season are also shifted. Another noticeable difference are the critical case-carrier ratio values (the lowest turning point in the epidemic region). These are smaller than those of the corresponding values for the deterministic model. All these suggest that there must be caution in assuming that the deterministic model approximates the stochastic mean equations.

**Fig. 4:**
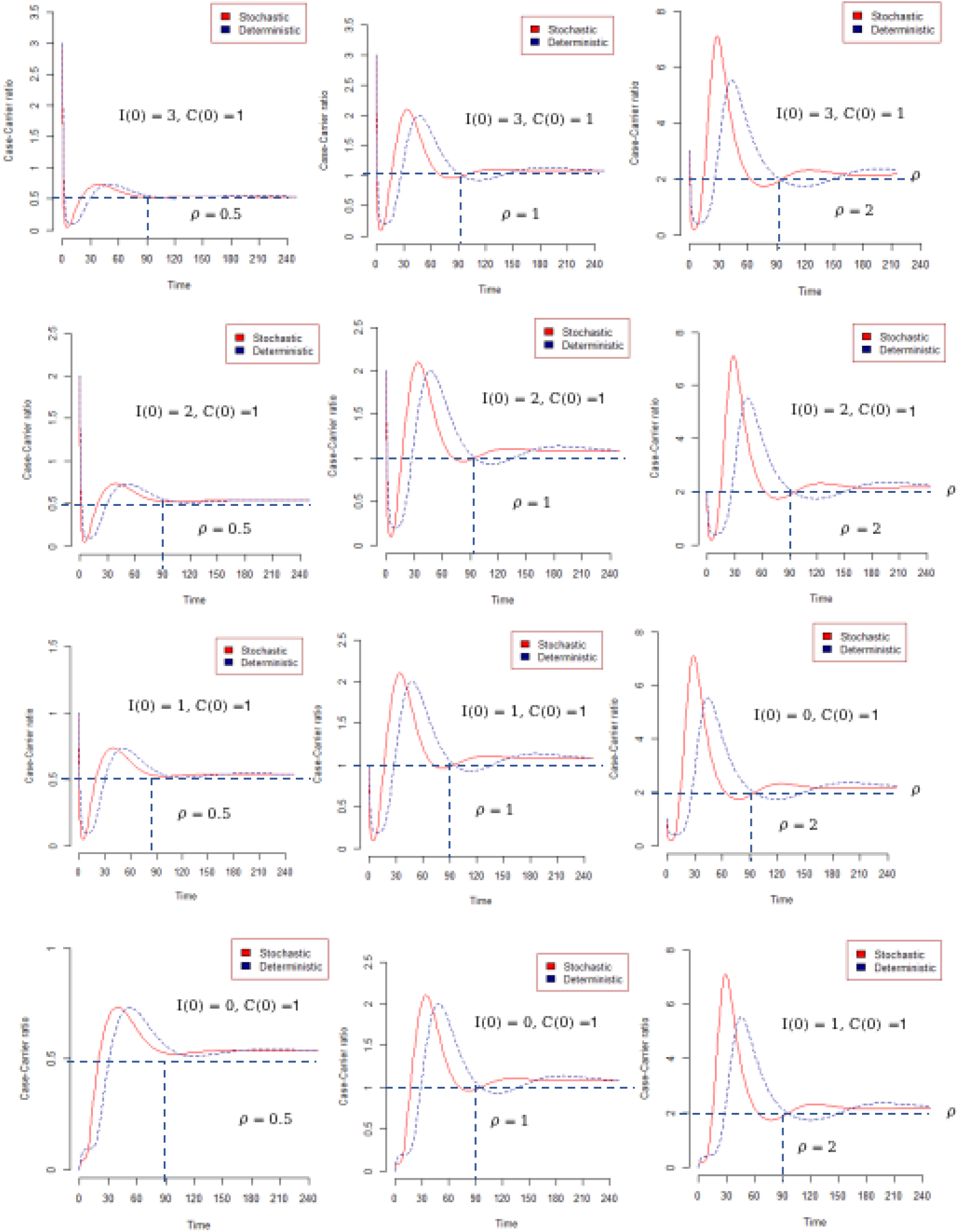
Case-Carrier ratio trajectory for stochastic means and deterministic model for various initial conditions and *ρ* = 0.5, 1 and *ρ* = 2

## 3. Simulation studies

### 3.1. Introduction

Generally, it is often difficult or mostly impossible to obtain an explicit expression that will help to explore some of the properties of a stochastic model describing an epidemic process. This is more true for meningitis epidemic process. However, to gain insight into the understanding of some of the salient features of stochastic epidemic process that will be of epidemiological importance requires the use of stochastic simulation (Dietz and Shenzle, 1985). Various attempts made for simulating stochastic epidemic process can be found in for example Whittle (1955), Ludwig (1973), Bailey (1975) and Keeling and Rohani (2008).The stochastic simulation allows one to numerically simulate the time evolution of a random system in a way that takes account of the randomness that is intrinsic in it and thus avoid mathematical intractability. Trajectories produced by simulation algorithms gives a more realistic representation of the system evolution than the conventional deterministic models (Gillespie, 1977 and 2001). Here we are using the Gillespie Tau-Leap simulation method. The foci are:

i. compare models I and II for various values of *ϵ* and
ii. see the effect of *ϵ* and *φ* on the carrier and infective incidences arising from models I and II.

### 3.2. Comparison of Models I and II for various values of *ϵ*

Figures 5 and 6 show the plots of the compartmental profiles for models I and II for values of *ϵ* = 0.5, 1 and 2 for *N* = 10^4^ and *N* = 5 *×* 10^4^ respectively. It is clear from these plots that as *ϵ* decreases in value below 2, the difference in models I and II becomes more pronounced for both sample sizes considered. However, when *ϵ* = 2, model II can approximate model I for all the compartmental profiles except that of the carrier. Here it is noticeable that the model I carrier profile peak value is considerably larger than that of model II.

**Fig. 5:**
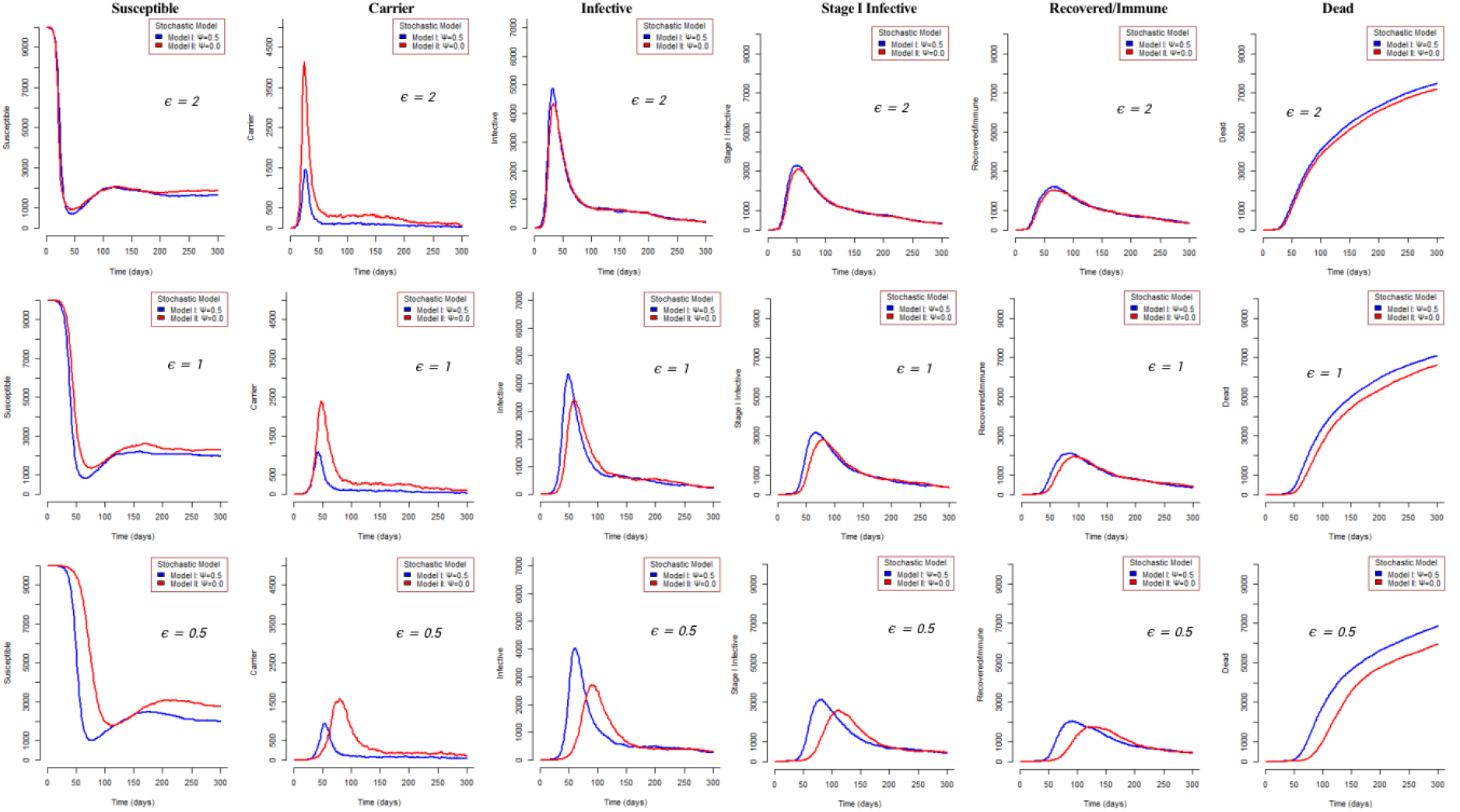
Plots of the compartmental profiles for models I and II for *ϵ* = 0.5, 1 and 2 for population size *N* = 10^4^

**Fig. 6:**
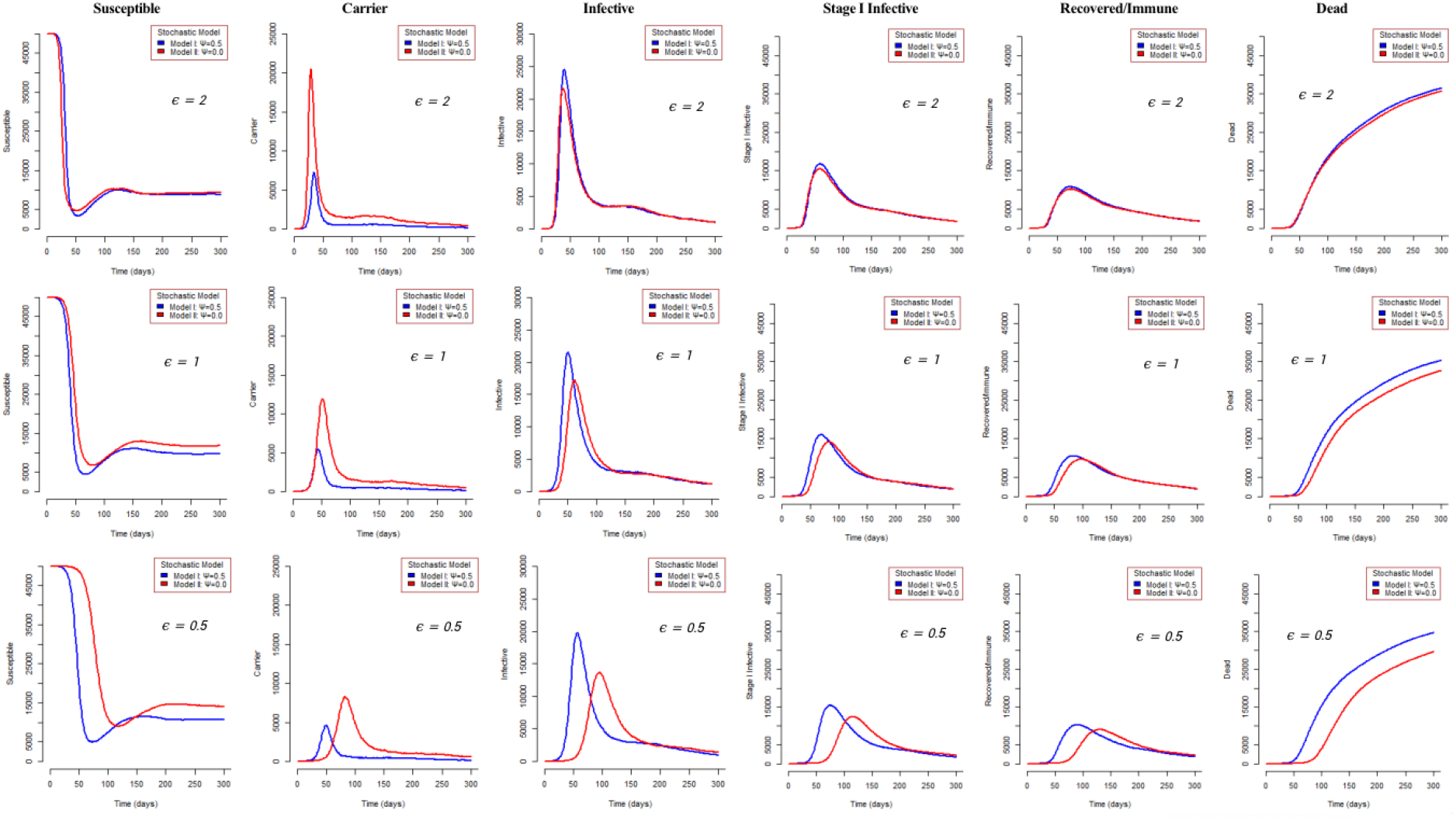
Plots of the compartmental profiles for models I and II for *ϵ* = 0.5, 1 and 2 for population size *N* = 5 *×* 10^4^

### 3.3. Effect of *ϵ* and *φ* on Carrier and Infective Incidence from models I and II

An important statistical tool that is employed by epidemiologists to understand the behaviour of a pathogen and its interaction with the host and its environment is the epidemic curve. The epidemic curve gives the rate at which new cases occur (Giesecke, 2002). Here we want to study the effect of *ϵ* = 0.5, 1, 2; *φ* = 0, 0.5 and *N* = 10^4^ and 5 *×* 10^4^ on the carrier and infective incidence curves for stochastic models I(*φ* = 0.5 and models II(*φ* = 0). The simulation is performed with initial conditions *C*(0) = 1 and *I*(0) = 1 and the plots are shown in Figs. 7-9. The corresponding deterministic curves are superimposed on these plots for the purpose of comparison. Also, estimates of important epidemiological characteristics of the infective and carrier incidences are shown in Tables 3-5 for ease of interpretation by the reader.

**Tab. 3:** Some values of the epidemiological characteristics of the infective incidence of meningitis epidemic derived from a stochastic model with initial conditions *C*(0) = 1, *I*(0) = 1 and population size *N* = 10, 000 and 50, 000.

| Stochastic |  |  |  |  |  |  |  |  |  |  |
| --- | --- | --- | --- | --- | --- | --- | --- | --- | --- | --- |
| Population size ($N$) | $\varphi$ | $\epsilon$ | Starting time of epidemic (${}_IT_1$) | End of epidemic time (${}_IT_2$) | Peak time of epidemic (${}_IT_{max}$) | Number of cases at peak $I({}_IT_{max})$ | Duration of epidemic (${}_IT_3$) | Total number recovered $R({}_IT_2)$ | Total Number of Death $D({}_IT_2)$ | Total number ever infected $n_I({}_IT_2)$ |
| 10,000 | 0.0 | 0.5 | 26 | 90 | 68 | 181 | 74 | 1087 | 414 | 2555 |
|  | 0.0 | 1.0 | 08 | 62 | 46 | 333 | 54 | 857 | 344 | 3314 |
|  | 0.0 | 2.0 | 06 | 40 | 28 | 726 | 34 | 851 | 228 | 4113 |
|  | 0.5 | 0.5 | 06 | 62 | 48 | 416 | 56 | 1221 | 527 | 3993 |
|  | 0.5 | 1.0 | 10 | 52 | 40 | 509 | 48 | 986 | 340 | 4166 |
|  | 0.5 | 2.0 | 04 | 42 | 30 | 663 | 38 | 336 | 277 | 4380 |
| 50,000 | 0.0 | 0.5 | 14 | 102 | 86 | 940 | 88 | 6487 | 2987 | 13374 |
|  | 0.0 | 1.0 | 04 | 60 | 48 | 2584 | 56 | 4529 | 2676 | 20933 |
|  | 0.0 | 2.0 | 06 | 44 | 34 | 3905 | 38 | 2288 | 1207 | 22118 |
|  | 0.5 | 0.5 | 08 | 72 | 58 | 1909 | 64 | 6368 | 2734 | 18607 |
|  | 0.5 | 1.0 | 04 | 60 | 48 | 2584 | 56 | 6139 | 2666 | 20933 |
|  | 0.5 | 2.0 | 04 | 46 | 36 | 4019 | 52 | 5578 | 1574 | 24542 |

**Fig. 7:**
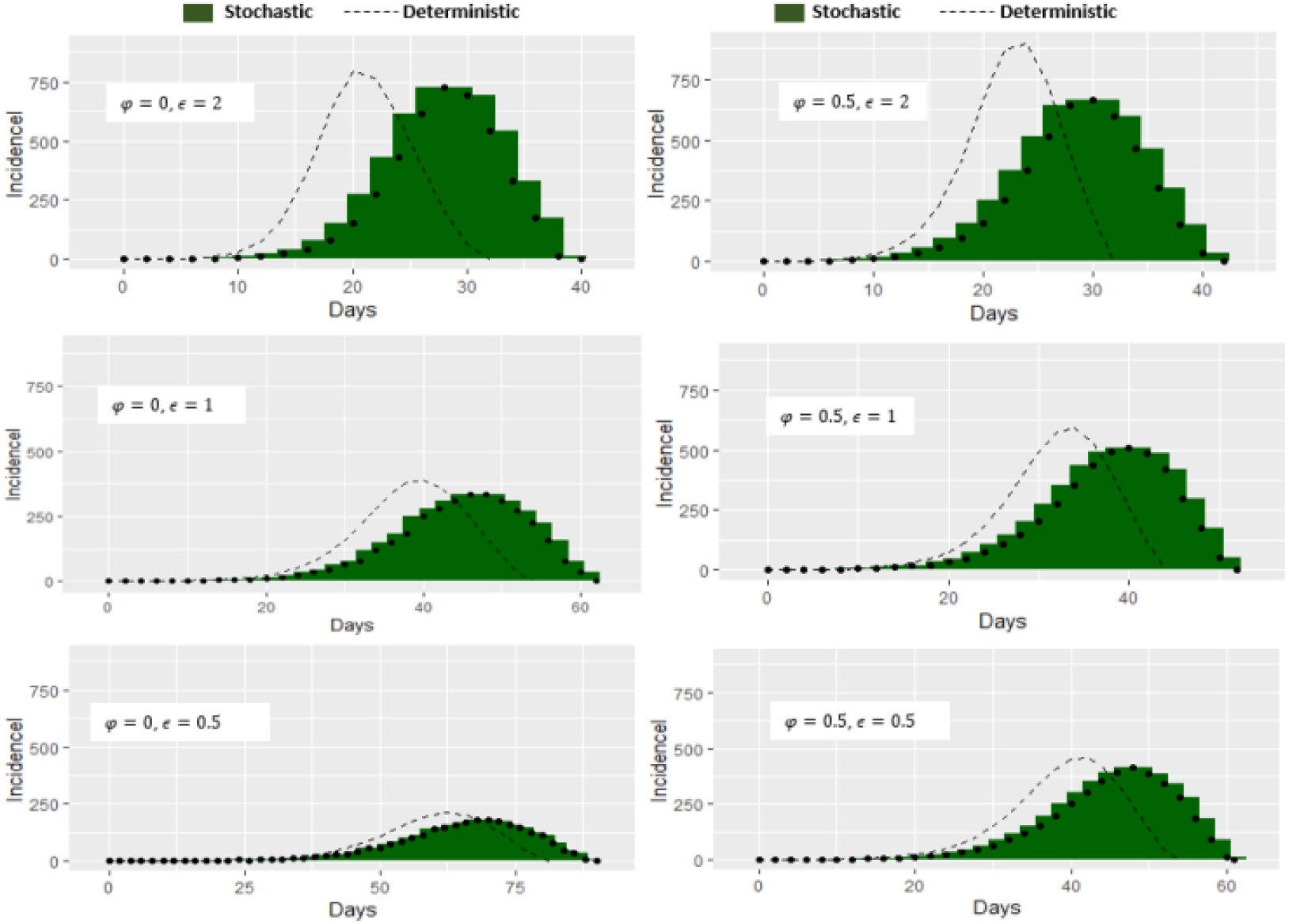
Epidemic Curves showing the effect of *ϵ* and *φ* for models I and Model II on infective incidence for population size *N* = 10^4^

#### 3.3.1. Infective Incidence Curve

Figures 7 and 8 show clearly that as the value of *ϵ* decreases so does that of the infective incidence, irrespective of *φ* and sample size. The same is also true for the deterministic model plots. However, there are noticeable differences in the stochastic and deterministic model plots that must be mentioned and are listed below with a provision for their actual values shown in Tables 3 and 4.

**Tab. 4:** Some values of the epidemiological characteristics of the infective incidence of meningitis epidemic derived from a deterministic model with initial conditions *C*(0) = 1, *I*(0) = 1 and population size *N* = 10, 000 and 50, 000.

| Deterministic |  |  |  |  |  |  |  |  |  |  |
| --- | --- | --- | --- | --- | --- | --- | --- | --- | --- | --- |
| Population size ($N$) | $\varphi$ | $\epsilon$ | Starting time of epidemic (${}_IT_1$) | End of epidemic time (${}_IT_2$) | Peak time of epidemic (${}_IT_{max}$) | Number of cases at peak $I({}_IT_{max})$ | Duration of epidemic (${}_IT_3$) | Total number recovered $R({}_IT_2)$ | Total Number of Death $D({}_IT_2)$ | Total number ever infected $n_I({}_IT_2)$ |
| 10,000 | 0.0 | 0.5 | 24 | 82 | 62 | 210 | 58 | 1072 | 645 | 2676 |
|  | 0.0 | 1.0 | 08 | 54 | 40 | 387 | 46 | 967 | 433 | 3314 |
|  | 0.0 | 2.0 | 06 | 32 | 20 | 791 | 26 | 870 | 285 | 4115 |
|  | 0.5 | 0.5 | 08 | 54 | 42 | 457 | 34 | 929 | 376 | 3821 |
|  | 0.5 | 1.0 | 08 | 44 | 34 | 591 | 26 | 951 | 353 | 4133 |
|  | 0.5 | 2.0 | 04 | 32 | 24 | 899 | 28 | 861 | 259 | 4678 |
| 50,000 | 0.0 | 0.5 | 16 | 94 | 76 | 1052 | 78 | 5320 | 3190 | 13383 |
|  | 0.0 | 1.0 | 10 | 62 | 46 | 1931 | 52 | 5074 | 2341 | 16559 |
|  | 0.0 | 2.0 | 06 | 36 | 24 | 4019 | 30 | 3884 | 1570 | 20549 |
|  | 0.5 | 0.5 | 06 | 62 | 50 | 2283 | 56 | 5310 | 2315 | 19108 |
|  | 0.5 | 1.0 | 06 | 52 | 40 | 2978 | 46 | 4161 | 2119 | 20704 |
|  | 0.5 | 2.0 | 04 | 38 | 28 | 4558 | 34 | 2501 | 1651 | 23425 |

**Fig. 8:**
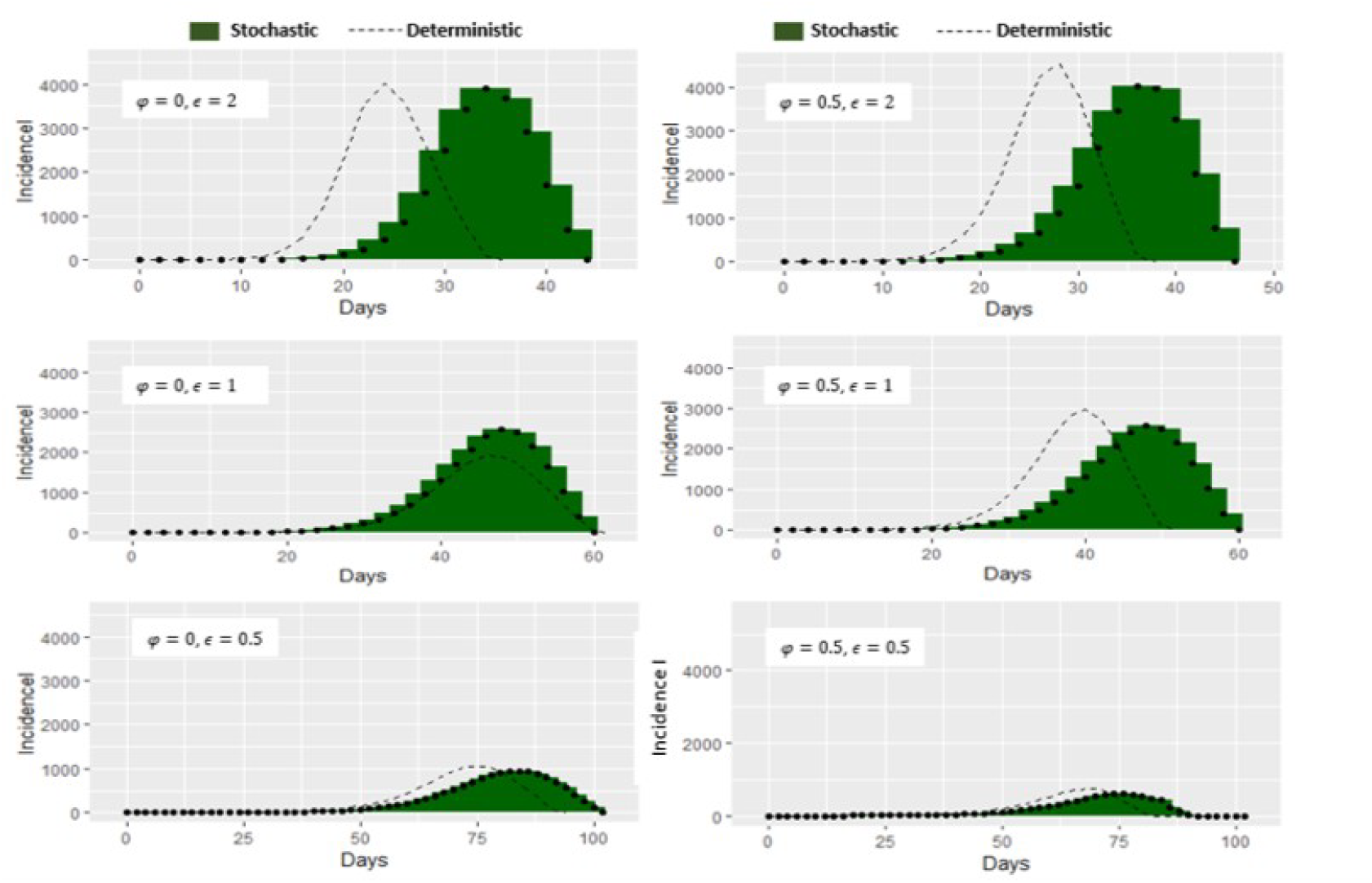
Epidemic Curves showing the effect of *ϵ* and *φ* for models I and Model II on infective incidence for population size *N* = 5 *×* 10^4^

1. The peak and stopping times of the infective incidence is earlier for the deterministic model.
2. The number of cases at the peak times is higher for the deterministic model.
3. It is only at the lowest value of *ϵ*(= 0.5) that the stochastic and deterministic model plots are reasonably close.

This indicates that in using the deterministic model as an approximating stochastic system, inferences emanating from such an assumption must carry an underlying precautionary note. Figures 7 and 8 show that as the value of *ϵ* becomes higher, plots for stochastic models show a trend of which model II(*φ* = 0) better approximate model I(*φ* = 0.5).

As can be seen in Tables 3 and 4, as the value of *ϵ* decreases, so also does the value of the total number ever infected for both the stochastic and the deterministic models. This identifies *ϵ* as an important parameter that needs to be reduced for any meaningful attempt at controling the spread of meningitis.

Epidemiological characteristics of the infective incidence of meningitis epidemic initiated by *C*(0) = 1 and *I*(0) = 1 for population size *N* = 10, 000 and *N* = 50, 000 are computed using the mean field of n=500 stochastic realization and the results shown in Table 3 below. The corresponding values for the deterministic model are shown in Table 4.

#### 3.3.2. Carrier Incidence

There are striking similarities and dissimilarities in the effect of *φ* and *ϵ* on the infective and carrier incidence as seen in Figures 7-10 and Tables 3-5. We shall be highlighting only differences here in order to avoid repetition. As seen in Figures 8 and 9, *φ* and *ϵ* both have influence on the carrier incidence. The effect of *ϵ* depends on *φ* particularly at high level of *ϵ*(= 2). This is true for both deterministic and stochastic plots. This is one of the major characteristic difference. It is also observable that as the value of *ϵ* decreases drastically:

**Fig. 9:**
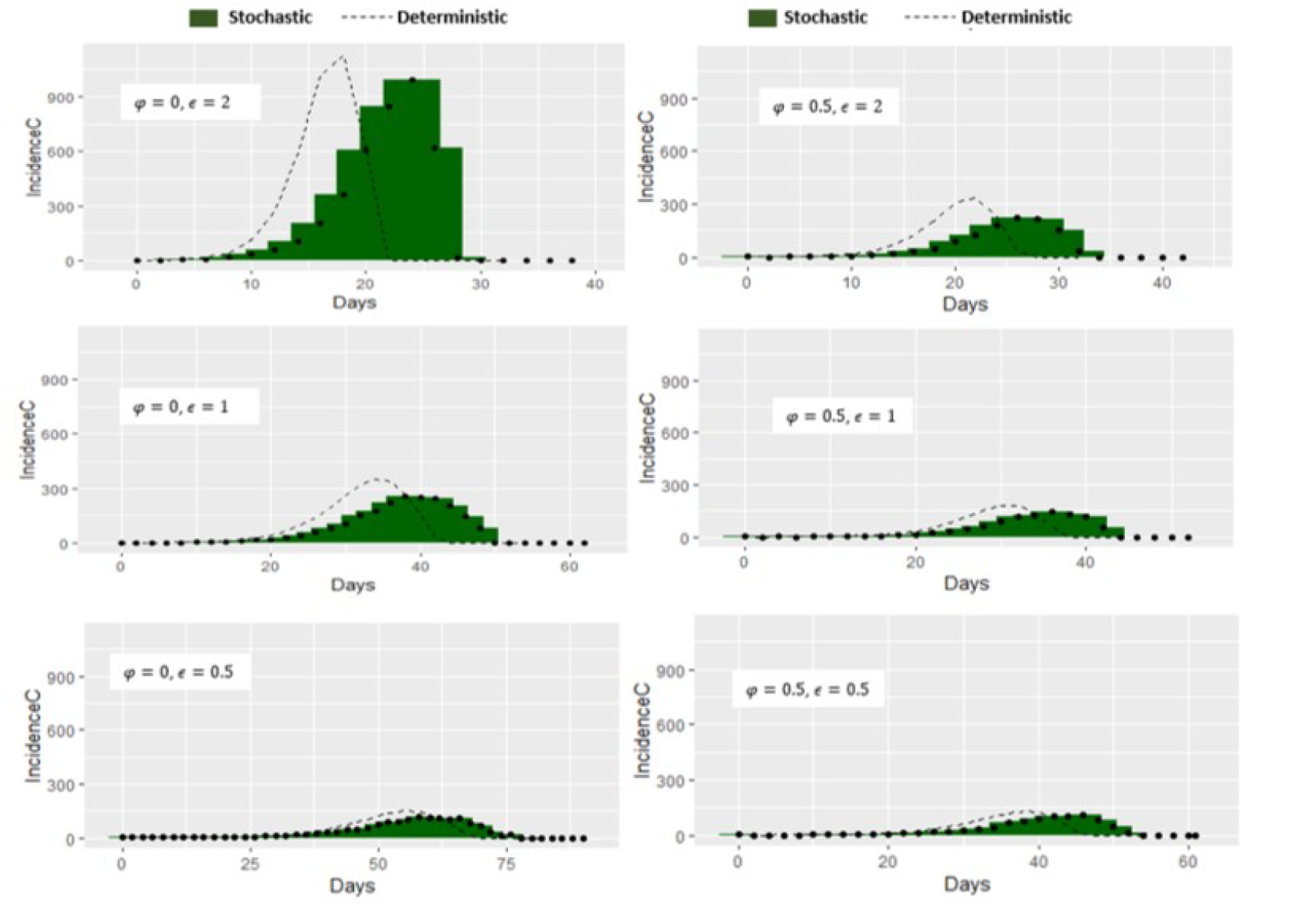
Epidemic Curves showing the effect of *ϵ* and *φ* for models I and Model II on Carrier incidence for population size *N* = 10^4^

**Fig. 10:**
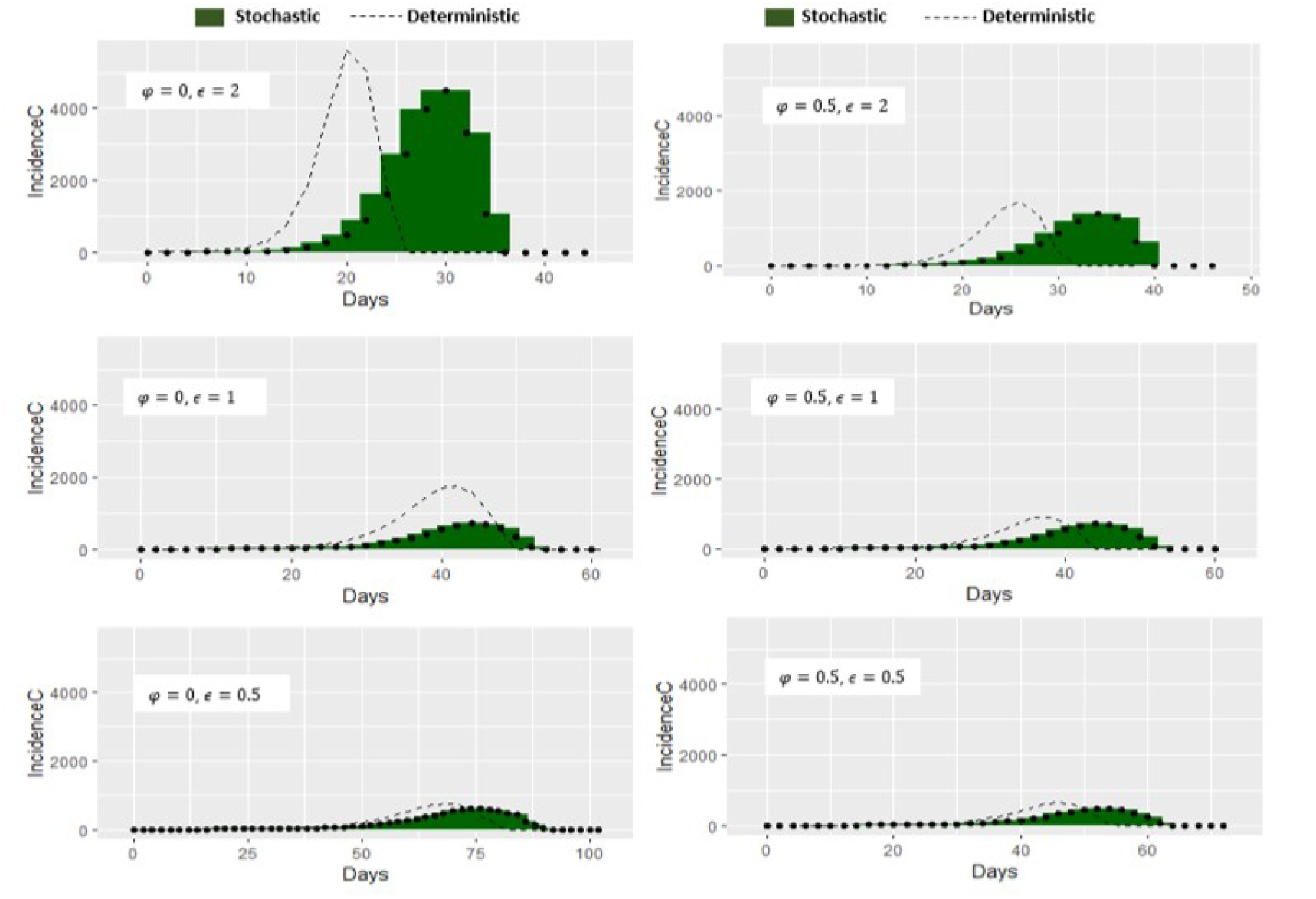
Epidemic Curves showing the effect of *ϵ* and *φ* for models I and Model II on Carrier incidence for population size *N* = 5 *×* 10^4^

i. influence of *ϵ* on the carrier plots drastically decreases irrespective of the value of *φ* and
ii. difference in the plots for the stochastic and deterministic models thin out and so also is the plots for model I (*φ* = 0.5) and model II (*φ* = 0).

These observations are also supported by corresponding values in Table 5. By implication

**Tab. 5:** Some values of epidemiological characteristics of Carrier incidence of meningitis epidemic derived from stochastic model with initial conditions *C*(0) = 1 and *I*(0) = 1 and population size *N* = 10, 000 and 50, 000

| Population size ($N$) | Stochastic | | | | | | Deterministic | | | |
| --- | --- | --- | --- | --- | --- | --- | --- | --- | --- | --- |
| | $\varphi$ | $\epsilon$ | Starting time of epidemic ($cT_1$) | End of epidemic time ($cT_2$) | Peak time of epidemic ($cT_{max}$) | Number of cases at peak $C(cT_{max})$ | Starting time of epidemic ($cT_1$) | End of epidemic time ($cT_2$) | Peak time of epidemic ($cT_{max}$) | Number of cases at peak $C(cT_{max})$ |
| $10^4$ | 0.0 | 0.5 | 16 | 77 | 58 | 117 | 16 | 73 | 56 | 150 |
|  | 0.0 | 1.0 | 08 | 50 | 38 | 254 | 12 | 45 | 30 | 179 |
|  | 0.0 | 2.0 | 02 | 28 | 24 | 854 | 02 | 22 | 18 | 1130 |
|  | 0.5 | 0.5 | 10 | 56 | 42 | 105 | 12 | 48 | 38 | 128 |
|  | 0.5 | 1.0 | 08 | 43 | 36 | 142 | 17 | 39 | 30 | 179 |
|  | 0.5 | 2.0 | 04 | 34 | 26 | 224 | 06 | 28 | 22 | 334 |
| $5 \times 10^4$ | 0.0 | 0.5 | 16 | 90 | 74 | 607 | 12 | 80 | 68 | 749 |
|  | 0.0 | 1.0 | 12 | 53 | 44 | 714 | 10 | 50 | 42 | 1761 |
|  | 0.0 | 2.0 | 04 | 36 | 30 | 4505 | 04 | 26 | 20 | 5608 |
|  | 0.5 | 0.5 | 14 | 62 | 54 | 483 | 10 | 55 | 46 | 640 |
|  | 0.5 | 1.0 | 08 | 53 | 44 | 714 | 10 | 45 | 38 | 903 |
|  | 0.5 | 2.0 | 08 | 36 | 34 | 1384 | 04 | 33 | 26 | 1693 |

a. the reduction of *ϵ* should be a target for meaningful control strategy for meningitis eradication as noted earlier for the infective case and
b. when *ϵ <* 1 the deterministic model can provide a first approximation to the stochastic model and model II (*φ* = 0) can be used as an approximating system of model I (*φ* = 0.5) in both the deterministic and stochastic situations.

Epidemiological characteristics of the carrier incidence of meningitis epidemic initiated by *C*(0) = 1 and *I*(0) = 1, for population size *N* = 10, 000 and 50, 000 are computed using the mean field of n=500 stochastic realizations and the results shown in Table 5 below. The corresponding values for deterministic model are also shown on the same table.

Again as in the infective case, Table 5 provide evidence that underlying caution should be borne in mind in making inferential statements emanating from the use of deterministic model in approximating its stochastic analog.

## 4. Stochastic Extinction of Meningitis epidemic

### 4.1. Introduction

Branching process is used as an approximate method for determining the probability of disease extinction or persistence, using the infectious classes with initial number of susceptible assumed to be at disease-free equilibrium points (Allen, 2012; Allen, Lahodny et al., 2015). The problems associated with approximating extinction probabilities are discussed in Britton et al. (2014). Here, the idea of Keeling and Rohani (2008) is used to derive the probability of disease extinction for meningitis epidemic.

Interest lies in studying the extinction probabilities of meningitis epidemic initiated by an infective and/or a carrier. In particular, we are using model II(*φ* = 0), previously mentioned to allow for mathematical tractability.

#### 4.1.1. Extinction of meningitis epidemics

##### Case1. Epidemic solely initiated by a carrier: *C*(0) = 1 and *I*(0) = 0

When a carrier is introduced in a population of susceptible with no infective in the population, the following are possible at the beginning of the process:

1. A carrier (C) can lose carriership with probability (*σ* + *φ* + *ϵβ*)^*−*1^*σ* and go to extinction with probability one
2. A carrier can convert to an infective with probability (*σ* + *φ* + *ϵβ*)^*−*1^*φ* and go into extinction with probability 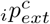.
3. A carrier can infect a susceptible to produce a carrier with probability (*σ* + *φ* + *ϵβ*)^*−*1^*ϵβ* which results in two carriers. The two carriers go into extinction with probability 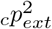.

The extinction probability for this epidemic process is obtained by summing all conditional probabilities in (i)-(iii) using the idea of Keeling and Rohani (2008), this is given by

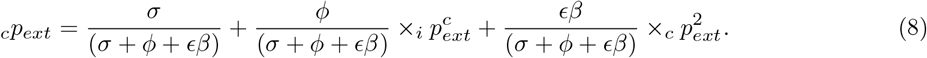

Equation (8) cannot be solved analytically due to the presence of 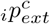. The notation 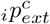 is adopted because it provides a leverage for anchoring some of the assumptions needed for mathematical tractability.

##### Case2. Epidemic solely initiated by an infective: *C*(0) = 0 and *I*(0) = 1

When an infective is assumed to be introduced in a population of susceptible, the following are possible at the beginning of the process:

i. The infective (I) can recover with probability (*β* + *θ* + *γ*_1_)^*−*1^*γ*_1_ and go into extinction with probability one.
ii. The infective can progress to stage one complication with probability (*β* + *θ* + *γ*_1_)^*−*1^*θ* and then go into extinction with probability one, for reason of being out of circulation.
iii. The infective can infect a susceptible with probability (*β* + *θ* + *γ*_1_)^*−*1*β*^ to produce one carrier; resulting in one carrier and one infective. The carrier goes into extinction with probability_*c*_*p*_*ext*_ and the infective goes to extinction with probability _*i*_*p*_*ext*_.

Summing all the conditional probabilities in Case2 (i)-(iii) the extinction probability for this epidemic process is given by

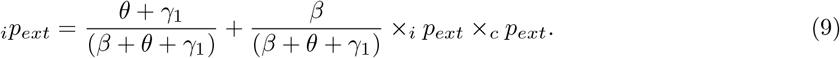

Equation (9) cannot be solved analytically. We make simplifying assumptions to make it tractable. We assume that the infected carrier will either convert to an infective with probability (*φ* + *σ*)^*−*1*φ*^ and go into extinction with probability *ip*_*ext*_ resulting in two infectives going into extinction with probability 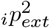 or an infected carrier can lose carriership with probability (*φ* + *σ*)^*−*1*σ*^ and go into extinction with probability one. This now results into only one infective going into extinction with probability _*i*_*p*_*ext*_. It must be mentioned that by allowing the converted carrier infective to go into extinction with probability _*i*_*p*_*ext*_ we have approximated 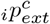 in case 1 by _*i*_*p*_*ext*_ for mathematical convenience. This is reasonable. Hence the extinction probability is now given by the equation

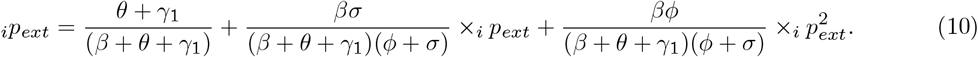

The solution for the probability of extinction for this equation is given by

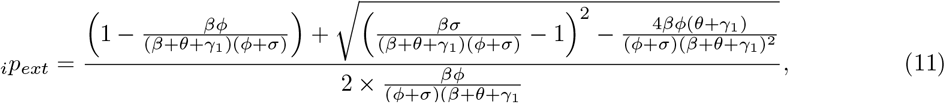

with condition for the real solution given by

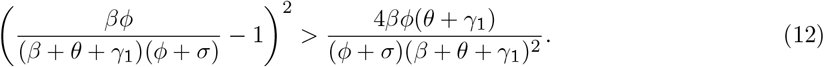

It is clear that not much can be deduced from this complex equation.

##### Case3(a). Epidemic initiated by one carrier and one infective: *C*(0) = 1 and *I*(0) = 1

###### Carrier Lineage

i. The carrier (C) can lose carriership with probability (*σ* + *φ* + *ϵβ*)^*−*1*σ*^ and go into extinction with probability one.
ii. The carrier can convert to an infective with probability (*σ* + *φ* + *ϵβ*)^*−*1*φ*^ and then go into extinction with probability 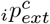.
iii. The carrier can infect a susceptible to produce a carrier with probability 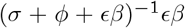 which resulting in two carriers. The two carriers go to extinction with probability 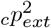.

###### Infective Lineage

i. The infective (I) can recover with probability (*β* + *θ* + *γ*_1_)^*−*1^_*γ*1_ and go into extinction with probability one.
ii. The infective can progress to stage one complication with probability (*β* + *θ* + *γ*_1_)^*−*1^*θ* and then go into extinction with probability one (due to being out of circulation).
iii. The infective can infect a susceptible with probability (*β* + *θ* + *γ*_1_)^*−*1^*β* resulting in one carrier and one infective. The carrier go to extinction with probability_*c*_*p*_*ext*_ and the infective goes to extinction with probability _*i*_*p*_*ext*_.

From the two formulations above,

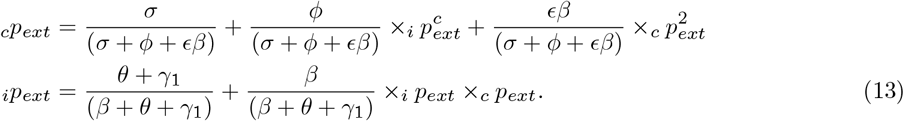

The system of equations (13) is nonlinear and hence non-tractable. Here as explained earlier we assumed that 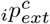, is approximated by _*i*_*p*_*ext*_ and equation (13) then becomes

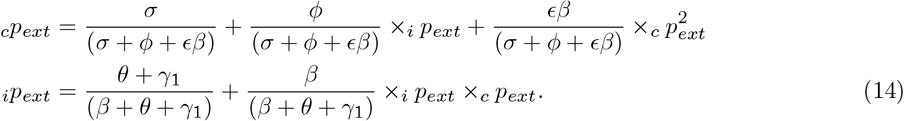

Eliminating _*i*_*p*_*ext*_ in (14), _*c*_*p*_*ext*_ becomes

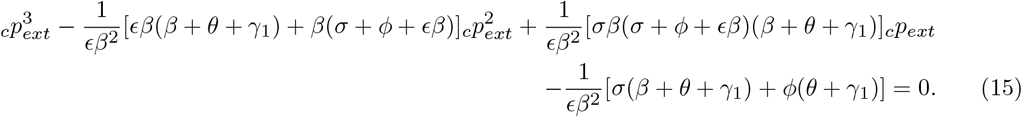

Equation (15) is a cubic equation. Using the Maple computer program (v.18) we obtained solutions with two complex and one real root. The real solutions for _*c*_*p*_*ext*_ and _*i*_*p*_*ext*_ are given by

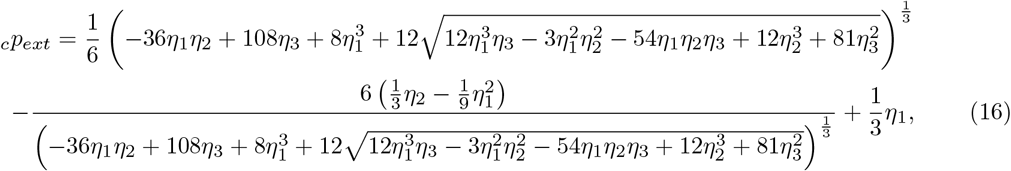

and

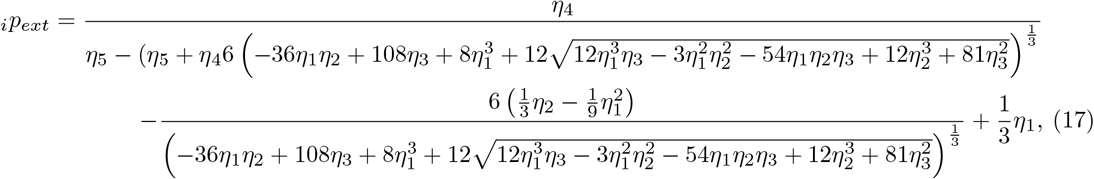

where

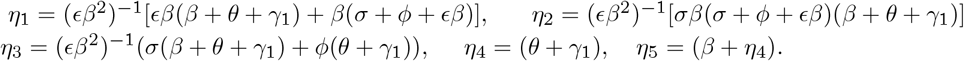

Equations (16) and (17) are cumbersome, and can hardly be used in throwing more light about the complex nature of extinction probabilities.

###### Case3(b). Epidemic initiated by one carrier and one infective: *C*(0) = 1 and *I*(0) = 1

In equation (7) we assume that the carrier converted infective goes into extinction to a first approximation with probability _*c*_*p*_*ext*_. That is 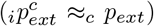. This is not far-fetched because 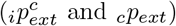 are all fractions and are likely not to be substantially different from one another, even at the first decimal place. This simplicity in assumption will make equation (18) amenable to mathematical exposition that can provide insight into the extinction probabilities of meningitis epidemic, which may be of importance for control intervention considerations.

Equation (7) then becomes

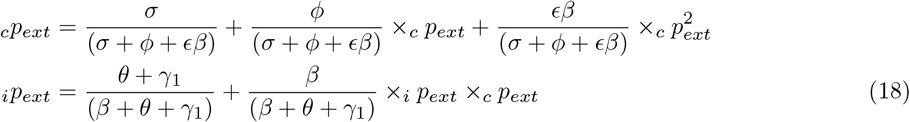

The solution to equation (18) is given by

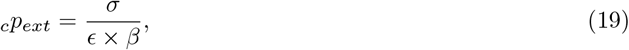

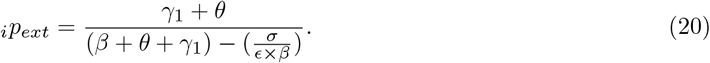

It must be mentioned that with these assumptions, the solution of equation (18) for _*c*_*p*_*ext*_ is now the same as for equation (8). Consequently deductions about _*c*_*p*_*ext*_ is also tenable for the initial condition *C*(0) = 1 and *I*(0) = 0.

From equation the solution given by equations (19) and (20), it is clear that the relationship between _*i*_*p*_*ext*_ and _*c*_*p*_*ext*_ is given by

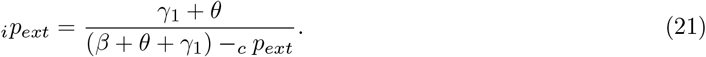

A graph of this for some parameter values is shown in Fig 11. This relationship is not our focus of discussion.

A condition on equation (19) is that 0 *≤* _*c*_*p*_*ext*_ *≤* 1, that is 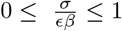. _*c*_*p*_*ext*_ = 0 only if *σ* = 0, that is, without loss of carriership _*c*_*p*_*ext*_ will be zero and hence there will be a spread of carriers. _*c*_*p*_*ext*_ = 1 if *σ* = *ϵβ*, implies that *ϵβ/σ* = 1. This can be interpreted by saying that the extinction of carriers is certain if the number of susceptibles infected by a carrier during the period of loss of carriership is one. If this number is greater than one, the carrier extinction is not certain. This is reasonable. Another interpretation can be obtained from

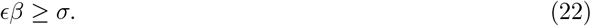

**Fig. 11:**
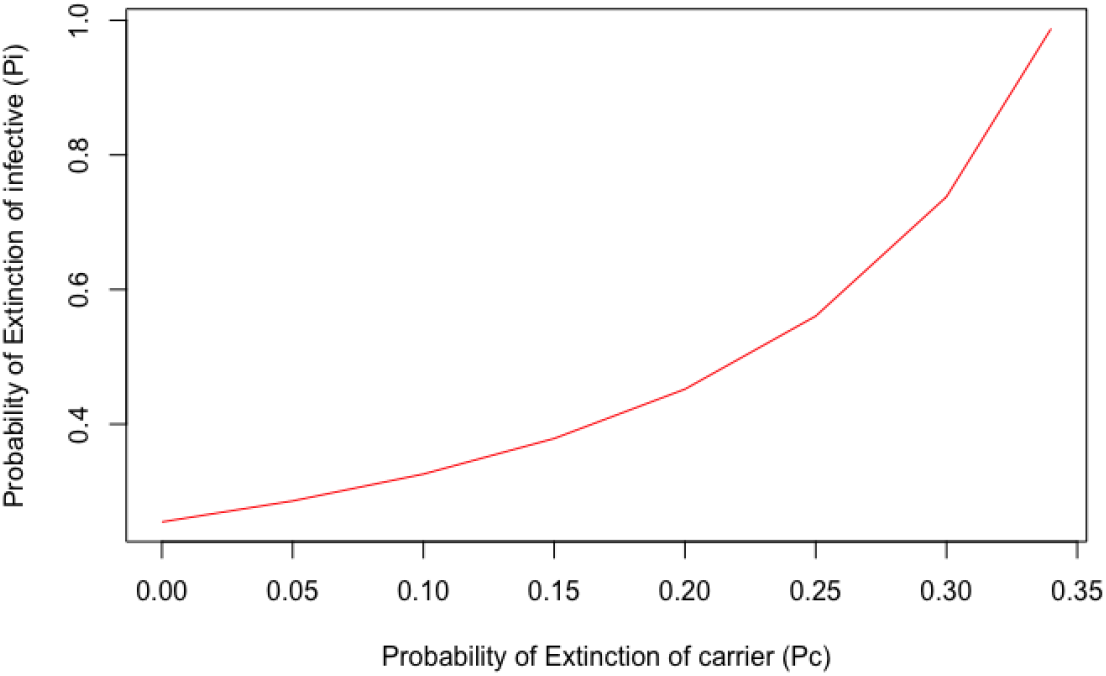
Relationship between _*i*_*p*_*ext*_ and _*c*_*p*_*ext*_ for some parameter values

This implies that the carrier transmission rate (*ϵβ*) has a threshold value *σ*, which is the carrier recovery rate for which the extinction of carrier is certain. It must be noted that the conditions are also true for epidemic initiated by *C*(0) = 1 and *I*(0) = 0 as explained earlier.

From equation (20), 0 *≤*_*i*_ *p*_*ext*_ *≤* 1, that is,

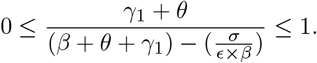

Hence, if 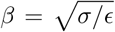, then _*i*_*p*_*ext*_ = 1 and then the extinction of infectives is certain. But if 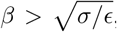, _*i*_*p*_*ext*_ *<* 1, then the extinction of infectives is not certain. Here

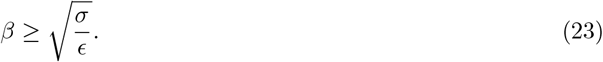

is a threshold equation which is quite different but related to that of the carrier. This means that if the infective transmission rate (*β*) equals a value 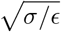 then the meningitis epidemic becomes extinct and if greater it persists.

It is worthy to note that *β, σ* and *ϵ* are the important epidemiological parameters that influence both the extinction of carriers and infectives.

### 4.2. Sensitivity Analysis

We are interested in computing the sensitivity indices of _*c*_*p*_*ext*_ and _*i*_*p*_*ext*_ to changes in the parameters of extinction probabilities. These indices give the relative contribution of the parameters and the nature of their influences on the extinction probability of meningitis epidemic. Consequently, they serve as a crucial information for any control strategies. The normalized forward sensitivity index of a variable to a parameter as defined in Yaga and Saporu (2024), and stated below is used for this computation. The sensitivity of the extinction probability, for example for _*c*_*p*_*ext*_ denoted by *S*_*c*_*p*_*ext*_, is derived from

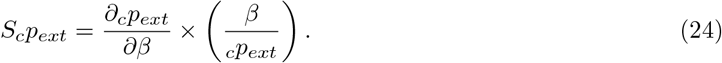

These indices are computed for _*c*_*p*_*ext*_ and _*i*_*p*_*ext*_ at specified parameter values from equations (11), (19) and (20) and are shown in Tables 6 to 8 respectively.

**Tab. 6:**
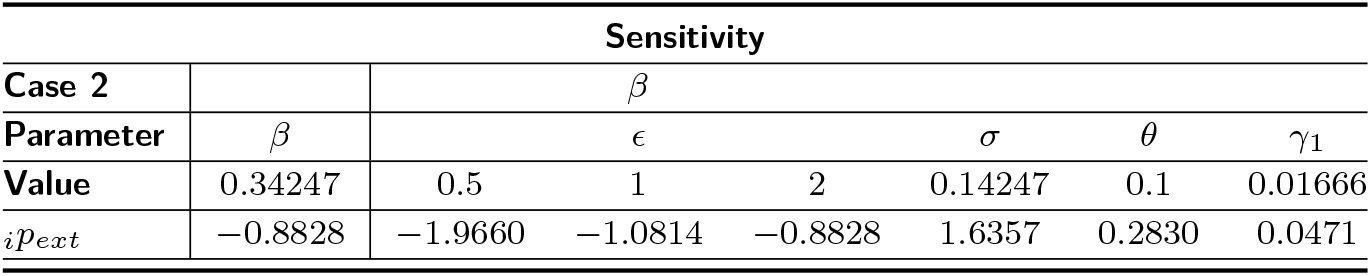
Sensitivity analysis for parameters of extinction probability (_*i*_*p*_*ext*_) for epidemic initiated by *C*(0) = 0 and *I*(0) = 1

**Tab. 7:**
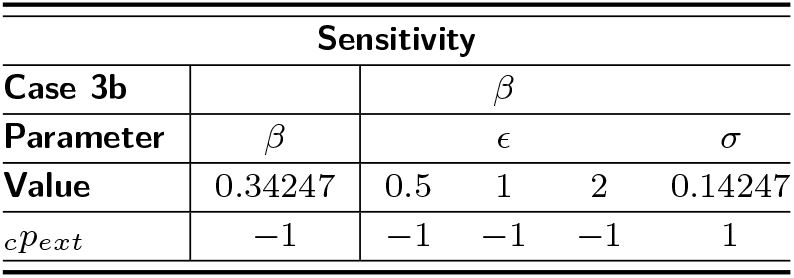
Sensitivity analysis for parameters of extinction probability (_*c*_*p*_*ext*_) for epidemic initiated by *C*(0) = 1 and *I*(0) = 1

**Tab. 8:** Sensitivity analysis for parameters of extinction probability (_*i*_*p*_*ext*_) for epidemic initiated by *C*(0) = 1 and *I*(0) = 1

| Sensitivity |  |  |  |  |
| --- | --- | --- | --- | --- |
| Case 3b |  |  |  |  |
| Parameter | $\beta$ | $\sigma$ | $\theta$ | $\gamma_1$ |
| Value | 0.34247 | 0.14247 | 0.1 | 0.01666 |
| $i p_{ext}$ | -0.5129 | 0.0040 | 0.1015 | 0.0338 |

The sensitivity indices are computed for scenarios where the extinction probability functions can be explicitly determined. Due to the complexity of some of the probability functions, the Maple computer program (Version 18) is used for these computation.

#### Discussion

From these tables, it is clear that the highest index values are obtained for parameters *β, ϵ* and *σ*, in almost all the cases considered. Values for *σ* are positive indicating that an increase in the value of *σ* will bring about a corresponding increase in the extinction probability under consideration. For *ϵ* and *β*, the values are negative, indicating that an increase in any of these parameter estimates will bring about a decrease in the extinction probability under consideration. This is as expected. Again this gives credence to the overriding influence of these parameters on extinction probabilities of meningitis epidemic

#### 4.2.1. Computing _*c*_*pext* and _*i*_*pext* for various parameters

Table 9 is computed using the analytic solutions obtained for case 3*b* (equations (19) and (20)) for various values of *β*(for *ϵ* = 2, 1, 0.5). It is clearly shown in Table 9 that an increase in the transmission rate of meningitis reduces the probability of disease extinction for _*c*_*p*_*ext*_ and _*i*_*p*_*ext*_.

**Tab. 9:** Extinction probabilities of carrier (_*c*_*p*_*ext*_) and infective (_*i*_*p*_*ext*_) for varying values of *β*, for *ϵ* = 0.5, 1, 2, *C*(0) = 1 and *I*(0) = 1

| Extinction Probability |  |  |  |  |  |  |
| --- | --- | --- | --- | --- | --- | --- |
| Parameter | $c p_{ext}$ | | | $i p_{ext}$ | | |
| | $\epsilon$ | | | $\epsilon$ | | |
| $\beta$ | 2 | 1 | 0.5 | 2 | 1 | 0.5 |
| 0.07120 | 1.00000 | - | - | 1.00000 | - | - |
| 0.07915 | 0.90000 | - | - | 0.93640 | - | - |
| 0.08904 | 0.80000 | - | - | 0.86757 | - | - |
| 0.10176 | 0.70000 | - | - | 0.79259 | - | - |
| 0.11873 | 0.60000 | - | - | 0.71235 | - | - |
| 0.14247 | 0.50000 | - | - | 0.62088 | - | - |
| 0.17809 | 0.40000 | 0.80000 | - | 0.52194 | 0.76610 | - |
| 0.23745 | 0.30000 | 0.60000 | - | 0.41241 | 0.55122 | - |
| 0.35618 | 0.20000 | 0.40000 | 0.80000 | 0.29049 | 0.35312 | 0.62087 |
| 0.71235 | 0.10000 | 0.20000 | 0.40000 | 0.15395 | 0.16992 | 0.21442 |
| 0.80000 | 0.08904 | 0.17809 | 0.35618 | 0.13799 | 0.15069 | 0.18467 |
| 0.90000 | 0.07915 | 0.15830 | 0.31660 | 0.12339 | 0.13344 | 0.15943 |
| 0.99999 | 0.07124 | 0.14247 | 0.28494 | 0.11159 | 0.11975 | 0.14026 |

Table 10 shows the comparison of _*c*_*p*_*ext*_ and _*i*_*p*_*ext*_ for various values of *σ* again using equation (19). These results show that an increase in the rate of loss of carriage can increase the probability of extinction of meningitis epidemic for both _*c*_*p*_*ext*_ and _*i*_*p*_*ext*_. From these results, it can be concluded any control measures aimed at reducing the transmission rate and increasing the loss of carriership rate will bring meningitis epidemic to an end.

**Tab. 10:** The Extinction probabilities of carrier (_*c*_*p*_*ext*_) and infective (_*i*_*p*_*ext*_) for varying values of *σ*, for *ϵ* = 2, *C*(0) = 1 and *I*(0) = 1

| Parameter | Extinction Probability |  |
| --- | --- | --- |
| $\sigma$ | $c_{P_{ext}}$ | $i_{P_{ext}}$ |
| 0.06849 | 0.10000 | 0.27457 |
| 0.13699 | 0.20000 | 0.29864 |
| 0.20548 | 0.30000 | 0.32734 |
| 0.27398 | 0.40000 | 0.36214 |
| 0.34247 | 0.50000 | 0.40522 |
| 0.41096 | 0.60000 | 0.45993 |
| 0.47946 | 0.70000 | 0.53172 |
| 0.54795 | 0.80000 | 0.63007 |
| 0.61645 | 0.90000 | 0.77309 |
| 0.68494 | 1.00000 | 1.00000 |

## 5. Conclusion

A stochastic version of the deterministic model for meningitis epidemic developed by Yaga and Saporu (2024) is studied. Its Kolmogorov forward system of differential equations is derived. So also are its moment generating function, stochastic mean system of equations and extinction probabilities. All these are new. A comparison of the system of stochastic mean equations and its deterministic analogue of profiles for the various compartments and the case-carrier trajectories show similar pattern with notable time shift difference. This suggest that there must be an underlying caution in using a deterministic model as an approximating system of its stochastic mean equations. This conclusion is again re-emphasized in the simulation studies of the effect of *ϵ* and *φ* on the carrier and infective incidence curves.

Simulation studies of the compartmental profiles for stochastic models I (*φ ≠* 0) and model II (*φ* = 0) for various values of *ϵ*(*≤* 2) indicates that only at *ϵ* = 2 can model II approximate model I for all profiles except that of the carrier; carrier profiles show noticeable difference in peak values.

The extinction probability studies suggest the following:

1. *ϵ, β* and *σ* are the most sensitive parameters for the carrier and infective extinction probabilities of meningitis; increase in *σ*(rate of loss of carriership) brings about corresponding increase in the extinction probability while increase in each of *ϵ* and *β* brings about corresponding decrease in extinction probability.
2. There are different thresholds conditions resulting from the carrier and infective extinction probabilities. Carrier extinction is certain if the number of susceptible infected by a carrier during the loss of carriership is one. If greater than one carriage persists. The extinction of infections is certain if the transmission rate, 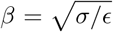. If 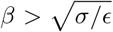, then the transmission of infection persists. Although these threshold conditions are distinct, they appear to be related. From equation (22), the transmission rate, *β*, required for carrier extinction is square that required for infective extinction. This lend credence to the thought that carriership play a more prominent role in the transmission process of meningitis epidemic (Borrow et al.2017; Irving et al.2012).
3. Control measures targeted at reducing the transmission rate and increasing the loss of carriership rate will erradicate meningitis epidemic. It should be noted here that similar conclusion was obtained from the deterministic model studies in Yaga and Saporu (2024).

## Data Availability

No Data used

